# Serum D-dimer level in women with polycystic ovary syndrome

**DOI:** 10.1101/2024.06.23.24308327

**Authors:** S Hossan, MS Morshed, H Banu, MA Hasanat

**Affiliations:** Department of Endocrinology, Bangabandhu Sheikh Mujib Medical University (BSMMU), Dhaka, Bangladesh; Department of Endocrinology, BSMMU, Dhaka, Bangladesh

**Keywords:** Polycystic ovary syndrome, D-dimer, Prothrombotic condition

## Abstract

**Background:** Women with polycystic ovarian syndrome (PCOS) have hyperandrogenemia and insulin resistance, which predispose them to prothrombotic conditions. D-dimer is a global marker of hemostatic dysfunction that has a contentious relationship with PCOS.

**Objective:** To assess the association of D-dimer with PCOS and its manifestations

**Methods:** This case-control study enrolled 44 women with PCOS based on the International Evidence-based Guideline, 2018, and an equal number of matched healthy controls by convenient sampling. After obtaining informed consent, participants’ clinical data was taken, and fasting blood was drawn to measure glucose, lipid profile, hormones, and D-dimer. D-dimer was analyzed by latex immunofluorescence assay.

**Results:** The D-dimer levels and status were statistically similar between the study groups [PCOS vs. control: 0.11 (0.10-0.17) vs. 0.13 (0.10-0.22), median (IQR), p= 0.673]. D-dimer levels did not vary according to the different characteristics of women with PCOS (p>0.05). D-dimer levels had no significant correlations with various clinical and biochemical characteristics among women with PCOS (p>0.05).

**Conclusion:** D-dimer has no significant association with PCOS or its manifestations.

## Introduction

Polycystic ovary syndrome (PCOS) is a common life-long problem in women with heterogeneous presentations.^1^ During their reproductive years, affected women typically report cutaneous and reproductive issues; later in life, they may experience cardiovascular morbidities.^2^ Atherosclerosis and inflammation-induced cardiovascular morbidities are exacerbated by changes in sex-steroid levels, increased insulin resistance, metabolic syndrome, and hemostatic abnormalities in women with PCOS.^3^ Abnormal platelet activation, endothelial dysfunction, hypercoagulability, and poor fibrinolysis are among the hemostatic abnormalities. Every one of these processes causes a prothrombotic condition in PCOS.^4^ Moreover, the first-line treatment for PCOS, combined oral contraceptives (COCs), increases the risk of venous thromboembolism.^5^ Women with PCOS are also more likely to have a family history of venous thrombosis.^6^ Furthermore, spontaneous abortion is a possible outcome of the prothrombotic condition.^7^ Therefore, for therapeutic management and cardiovascular disease prevention, the hemostatic assessment in women with PCOS is crucial.

The clotting cascade is the basic component of the blood coagulation process. It includes both coagulation and fibrinolysis, which result in blood clot production and removal from the system. The blood coagulation cascade involves the production of clots and their disposal from the system via two distinct paths known as the intrinsic and extrinsic pathways. The intrinsic process begins with the activation of blood clotting factors XI and XII, whereas the extrinsic pathway begins with the production of tissue factor (TF), which leads to the activation of Factor X. These two processes eventually lead to the development of fibrin (clot), which is degraded by plasmin into D-dimer.^8,9^ Its measurement evaluates the fibrinolytic and coagulation systems comprehensively. Conflicting findings from earlier research linked D-dimer to PCOS and its symptoms. Since our race predominates metabolic phenotypes, evaluating the connection would be pertinent.^10^ Our study’s goals were to evaluate the relationship between PCOS and D-dimer in reproductive-aged women.

## Materials & Methods

This case-control study was done in the Department of Endocrinology, Bangabandhu Sheikh Mujib Medical University (BSMMU) between September 2020 and August 2022. The Institutional Review Board of BSMMU (R/N no: 3582; BSMMU/2021/7642, date: 24/08/2021) provided the ethical clearance for the study. Informed consent was taken from each participant.

Considering the following formula [n= (Z_α_+Z_β_)^2^×(_σ1_^2^+_σ2_^2^) ÷ (μ_1_-μ_2)_^2^], using the mean and SD (PCOS: 280.6±69.4, control: 227.6±73.9, ng/mL) from a previous study, and considering 95% confidence interval and 90% power, the sample size was 38.^11^ We enrolled 44 women aged 18 – 35 years with PCOS and an equal number of matched healthy controls. We diagnosed PCOS according to the International Evidence-based Guideline, 2018.^12^ Healthy controls were those who had regular cycles, no hyperandrogenism symptoms, and normal ovarian morphology. Everyone who had comparable endocrinopathies, severe and uncontrolled systemic illnesses, a history of COVID-19 within the last four months, had used aspirin, COCs, antiandrogens, insulin sensitizers, or an anticoagulant within the previous six months, or who was pregnant or breastfeeding was excluded. After taking relevant clinical information from PCOS women, fasting blood was drawn to measure glucose, insulin, and D-dimer. Glucose was measured by glucose oxidase, insulin by chemiluminescence immunoassay, D-dimer by latex immunofluorescence assay by Getein 1100 immunofluorescence quantitative analyzer, Getein Biotech, Inc., China. The minimum detectable level was 0.1 mg/L with an inter- and intra-assay coefficient of variation of below 10%. Insulin resistance was calculated from homeostatic model assessment (HOMA) and a cut-off of 2.6 was set.^13^ A D-dimer level below 0.5 mg/L was considered normal.^14^

Version 25.0 of SPSS software was used to analyze the data. Qualitative factors were expressed in frequency (%), whereas quantitative data were expressed in mean ± SD or median (IQR) depending on their distributions. Fisher’s exact test was used to analyze the qualitative variables and the Student’s t-test or Mann-Whitney U test was used to analyze the associations between various quantitative variables and PCOS or its symptoms. The Spearman’s correlation test was used to see whether D-dimer and certain PCOS characteristics were correlated. Any two-tailed p-value below 0.05 was considered statistically significant.

## Results

The characteristics of the study participants showed that women with PCOS had poorer metabolic profiles (higher BMI, systolic & diastolic blood pressure, waist circumference, and HOMA-IR) than healthy control. The D-dimer levels and status were statistically similar between the study groups (Table-I). D-dimer levels did not vary according to the different characteristics of women with PCOS (Table-II). D-dimer levels had no significant correlations with different clinical and biochemical characteristics among women with PCOS (Table-III).

**Table-I:**
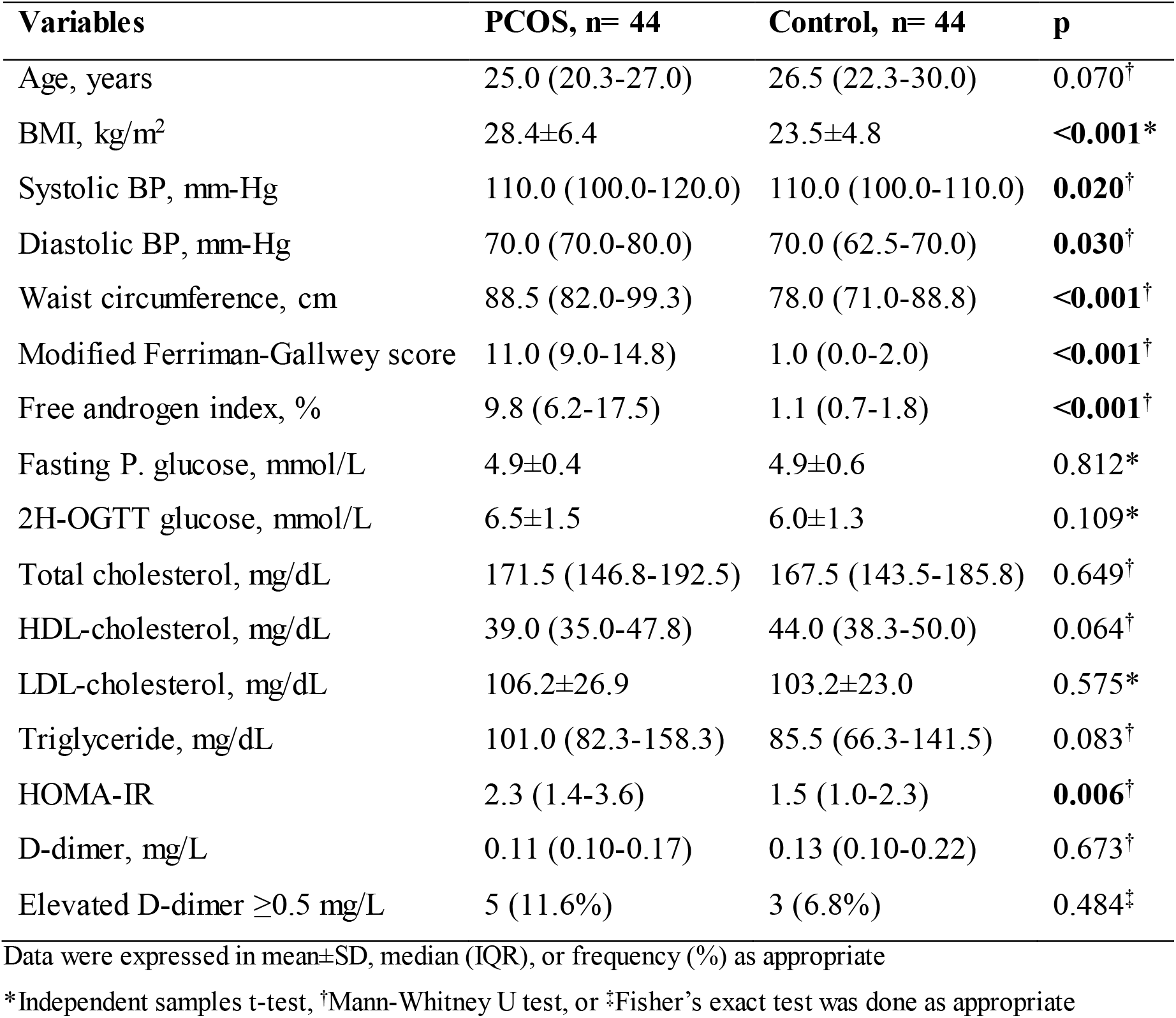
Characteristics of the study participants (n= 88)

**Table-II:** D-dimer levels with different characteristics of women with PCOS (n=44)

| Variables | Subgroups | No. | D-dimer levels, mg/L | p |
| --- | --- | --- | --- | --- |
| Body mass index | Lean (<23 kg/m<sup>2</sup>) | 11 | 0.10 (0.10-0.11) | 0.301 |
|  | Overweight/obese (≥23 kg/m<sup>2</sup>) | 33 | 0.11 (0.10-0.19) |  |
| Biochemical hyperandrogenism | Absent (FAI <5%) | 8 | 0.11 (0.10-0.20) | 0.915 |
|  | Present (≥5%) | 36 | 0.11 (0.10-0.17) |  |
| Glycemic status | Normal glucose tolerance | 32 | 0.10 (0.10-0.19) | 0.968 |
|  | Abnormal glucose tolerance | 12 | 0.11 (0.10-0.15) |  |
| Insulin resistance | Absent (HOMA-IR <2.6) | 26 | 0.10 (0.10-0.16) | 0.366 |
|  | Present (≥2.6) | 18 | 0.11 (0.10-0.22) |  |
| Metabolic syndrome | Absent (<3/5 criteria) | 34 | 0.11 (0.10-0.15) | 0.505 |
|  | Present (≥3/5) | 10 | 0.12 (0.10-0.63) |  |
| LH/FSH ratio | Normal (≤2) | 25 | 0.11 (0.10-0.18) | 0.914 |
|  | Altered (>2) | 19 | 0.10 (0.10-0.17) |  |
| Polycystic ovarian morphology | Absent | 13 | 0.10 (0.10-0.32) | 0.744 |
|  | Present | 31 | 0.11 (0.10-0.17) |  |

Mann-Whitney U test was done

**Tab-III:** Correlation between D-dimer and different characteristics among women with PCOS (n=44)

Mann-Whitney U test was done **Tab-III:** Correlation between D-dimer and different characteristics among women with PCOS (n=44)
| **Variables** | **r<sub>s</sub>** | **p** |
| --- | --- | --- |
| Age, years | 0.17 | 0.268 |
| BMI, kg/m<sup>2</sup> | 0.16 | 0.315 |
| Waist circumference, cm | 0.18 | 0.256 |
| Systolic BP, mm-Hg | 0.21 | 0.178 |
| Diastolic BP, mm-Hg | 0.15 | 0.326 |
| Modified Ferriman-Gallwey score | -0.28 | 0.062 |
| Free androgen index, % | -0.13 | 0.417 |
| Fasting P. glucose, mmol/L | 0.14 | 0.360 |
| 2H-OGTT glucose, mmol/L | 0.03 | 0.833 |
| Total cholesterol, mg/dL | 0.04 | 0.815 |
| HDL-cholesterol, mg/dL | 0.001 | 0.995 |
| LDL-cholesterol, mg/dL | 0.01 | 0.928 |
| Triglyceride, mg/dL | -0.02 | 0.879 |
| LH/FSH ratio | -0.06 | 0.705 |
| HOMA-IR | 0.15 | 0.321 |

Spearman's correlation test was done

## Discussion

Women with PCOS possess several metabolic risk factors and hyperandrogenism that predispose them to a prothrombotic state. Measuring D-dimer levels may provide a clue about the patient’s risk of thrombosis especially while initiating COCs.^15^ In this context, we measure the D-dimer levels to see its association with PCOS and its characteristics. However, we found no association of D-dimer with PCOS and its different characteristics.

We found similar levels of D-dimer between PCOS and healthy control. The findings regarding the connection between PCOS and D-dimer levels are contradictory. Several investigations revealed a similar conclusion.^16-18^ Others, however, found that women with PCOS had higher levels of D-dimer than a healthy control group.^11,19,20^ Higher D-dimer levels suggest increased subclinical fibrin synthesis rather than a breakdown in the setting of decreased global fibrinolytic activity.^21^ While some researchers discovered no association at all or an association that varied depending on age and BMI, others discovered an independent positive association between D-dimer and PCOS.^22,23^ The prevalence of elevated D-dimer status was higher in our study than in the control group, although the differences were not statistically significant. We could not find any research that contrasts the frequency of high D-dimer status between PCOS patients and healthy controls. The absence of thrombotic characteristics in individuals with elevated D-dimer levels could potentially be attributed to a low grade of chronic inflammation.^24^ However, we did not measure any markers of inflammation in this indexed study.

We did not find any association or correlation between D-dimer levels and several features of PCOS. The association between D-dimer levels and different characteristics of PCOS is not evaluated adequately in the literature. Kebapcillar et al. (2009) found a positive correlation between D-dimer levels with HOMA-IR and free testosterone levels.^19^ However, the association remained significant only with free testosterone in a linear regression model. Similarly, Moin et al., (2021) found an insignificant association of D-dimer levels with BMI and HOMA-IR.^25^

The cross-sectional design and single-center tertiary care sample collecting site of this study are its limitations. Besides, we could not measure other markers of coagulation and fibrinolytic pathways as well as the markers of inflammation.

In summary, there is no discernible association between D-dimer and PCOS or its many manifestations. To determine the precise disruptions in the hemostatic system in PCOS-affected women, prospective studies may be recommended to evaluate additional hemostatic parameters and their correlation with thrombotic status.

## Data Availability

All data produced in the present study are available upon reasonable request to the authors

## Declarations

### Ethical approval

IRB, BSMMU

## Acknowledgment

Department of Microbiology & Immunology, BSMMU

## References

1. Che Y, Yu J, Li YS, Zhu YC, Tao T. Polycystic ovary syndrome: Challenges and possible solutions. J Clin Med 2023;12(4):1500. DOI: 10.3390/jcm12041500.

2. de Melo AS, Dias SV, Cavalli Rde C, Cardoso VC, Bettiol H, Barbieri MA, et al. Pathogenesis of polycystic ovary syndrome: Multifactorial assessment from the foetal stage to menopause. Reproduction 2015;150(1):R11–24. DOI: 10.1530/REP-14-0499.

3. Duică F, Dănilă CA, Boboc AE, Antoniadis P, Condrat CE, Onciul S, et al. Impact of increased oxidative stress on cardiovascular diseases in women with polycystic ovary syndrome. Front Endocrinol (Lausanne) 2021;12:614679. DOI: 10.3389/fendo.2021.614679.

4. Targher G, Zoppini G, Bonora E, Moghetti P. Hemostatic and fibrinolytic abnormalities in polycystic ovary syndrome. Semin Thromb Hemost 2014;40(5):600–18. DOI: 10.1055/s-0034-1384512.

5. Dragoman MV, Tepper NK, Fu R, Curtis KM, Chou R, Gaffield ME. A systematic review and meta-analysis of venous thrombosis risk among users of combined oral contraception. Int J Gynaecol Obstet 2018;141(3):287–94. DOI: 10.1002/ijgo.12455.

6. Atiomo WU, Fox R, Condon JE, Shaw S, Friend J, Prentice AG, et al. Raised plasminogen activator inhibitor-1 (PAI-1) is not an independent risk factor in the polycystic ovary syndrome (PCOS). Clin Endocrinol (Oxf) 2000;52(4):487–92. DOI: 10.1046/j.1365-2265.2000.00946.x.

7. Cavalcante MB, Sarno M, Cavalcante CTMB, Araujo Júnior E, Barini R. Coagulation biomarkers in women with recurrent miscarriage and polycystic ovarian syndrome: Systematic review and meta-analysis. Geburtshilfe Frauenheilkd 2019;79(7):697–704. DOI: 10.1055/a-0884-3212.

8. Chaudhry R, Usama SM, Babiker HM. Physiology, Coagulation Pathways. [Updated 2023 Aug 28]. In: StatPearls [Internet]. Treasure Island (FL): StatPearls Publishing; 2024 Jan-. Available from: https://www.ncbi.nlm.nih.gov/books/NBK482253/

9. Wilhelm G, Mertowska P, Mertowski S, Przysucha A, Strużyna J, Grywalska E, et al. The crossroads of the coagulation system and the immune system: Interactions and connections. Int J Mol Sci 2023;24(16):12563. DOI: 10.3390/ijms241612563.

10. Sendur SN, Yildiz BO. Influence of ethnicity on different aspects of polycystic ovary syndrome: A systematic review. Reprod Biomed Online 2021;42(4):799–818. DOI: 10.1016/j.rbmo.2020.12.006.

11. Oral B, Mermi B, Dilek M, Alanoğlu G, Sütçü R. Thrombin activatable fibrinolysis inhibitor and other hemostatic parameters in patients with polycystic ovary syndrome. Gynecol Endocrinol 2009;25(2):110–6. DOI: 10.1080/09513590802549874.

12. Teede HJ, Misso ML, Costello MF, Dokras A, Laven J, Moran L, et al. Recommendations from the international evidence-based guideline for the assessment and management of polycystic ovary syndrome. Hum Reprod 2018;33(9):1602–18. DOI: 10.1093/humrep/dey256.

13. Bhowmik B, Siddiquee T, Mujumder A, Rajib MMR, Das CK, Khan MI, et al. Identifying insulin resistance by fasting blood samples in Bangladeshi population with normal blood glucose. J Diabetol 2016;7:p4.

14. Bounds EJ, Kok SJ. D Dimer. [Updated 2023 Aug 31]. In: StatPearls [Internet]. Treasure Island (FL): StatPearls Publishing; 2024 Jan. Available from: https://www.ncbi.nlm.nih.gov/books/NBK431064/

15. Gariani K, Hugon-Rodin J, Philippe J, Righini M, Blondon M. Association between polycystic ovary syndrome and venous thromboembolism: A systematic review and meta-analysis. Thromb Res 2020;185:102–108. DOI: 10.1016/j.thromres.2019.11.019.

16. Demirpence M, Yilmaz Yasar H, Karakoyun I, Girgin EM. Galanin-like peptide and its correlation with androgen levels in patients with polycystic ovary syndrome. Endokrynol Pol 2023;74(2):197–202. DOI: 10.5603/EP.a2023.0026.

17. de Mendonça-Louzeiro MR, Annichino-Bizzacchi JM, Magna LA, Quaino SK, Benetti-Pinto CL. Faster thrombin generation in women with polycystic ovary syndrome compared with healthy controls matched for age and body mass index. Fertil Steril 2013;99(6):1786–90. DOI: 10.1016/j.fertnstert.2013.01.105.

18. Karakurt F, Gumus II, Bavbek N, Kargili A, Koca C, Selcoki Y, Ozbek M, Kosar A, Akcay A. Increased thrombin-activatable fibrinolysis inhibitor antigen levels as a clue for prothrombotic state in polycystic ovary syndrome. Gynecol Endocrinol 2008;24(9):491–97. DOI: 10.1080/09513590802291824.

19. Kebapcilar L, Taner CE, Kebapcilar AG, Sari I. High mean platelet volume, low-grade systemic coagulation and fibrinolytic activation are associated with androgen and insulin levels in polycystic ovary syndrome. Arch Gynecol Obstet 2009;280(2):187–93. DOI: 10.1007/s00404-008-0884-0.

20. Sun Q, Yang Y, Peng X, Zhang Y, Gao Y, Wang F, Zhang Y, Feng W, Yang W, Kang X. Coagulation parameters predictive of polycystic ovary syndrome. Eur J Obstet Gynecol Reprod Biol 2019;240:36–40. DOI: 10.1016/j.ejogrb.2019.06.018.

21. Yildiz BO, Haznedaroğlu IC, Kirazli S, Bayraktar M. Global fibrinolytic capacity is decreased in polycystic ovary syndrome, suggesting a prothrombotic state. J Clin Endocrinol Metab 2002;87(8):3871–75. DOI: 10.1210/jcem.87.8.8716.

22. Moin ASM, Sathyapalan T, Butler AE, Atkin SL. Coagulation factor dysregulation in polycystic ovary syndrome is an epiphenomenon of obesity. Clin Endocrinol (Oxf) 2023;98(6):796–802. DOI: 10.1111/cen.14904.

23. Mannerås-Holm L, Baghaei F, Holm G, Janson PO, Ohlsson C, Lönn M, Stener-Victorin E. Coagulation and fibrinolytic disturbances in women with polycystic ovary syndrome. J Clin Endocrinol Metab 2011;96(4):1068–76. DOI: 10.1210/jc.2010-2279.

24. Begum A, Morshed MS, Banu H, Zamila BM, Shah S, Hasanat MA. Association of high sensitivity C-reactive protein in polycystic ovary syndrome. J Dhaka Med Coll 2021;29(2):142–48. DOI: 10.3329/jdmc.u30i2.5691B.

25. Moin ASM, Sathyapalan T, Diboun I, Elrayess MA, Butler AE, Atkin SL. Metabolic consequences of obesity on the hypercoagulable state of polycystic ovary syndrome. Sci Rep 2021;11(1):5320. DOI: 10.1038/s41598-021-84586-y.

